# Estimating vaccine confidence levels among healthcare staff and students of a tertiary institution in South Africa

**DOI:** 10.1101/2021.09.17.21263739

**Authors:** Elizabeth O. Oduwole, Tonya Esterhuizen, Hassan Mahomed, Charles S. Wiysonge

## Abstract

**Introduction:** In South Africa, healthcare workers were the first group scheduled to receive a COVID-19 vaccine when it became available. Therefore, estimating their vaccine confidence levels and gauging their intention to vaccinate ahead of the COVID-19 vaccination roll-out was imperative.

**Methods:** An online survey was conducted among current staff and students of the Faculty of Medicine and Health Sciences of Stellenbosch University in South Africa using a succinct questionnaire. Sentiments about vaccines were estimated using five validated statements. The intention to receive a COVID-19 vaccine was also investigated.

**Results:** The response rate was 21.8%, giving a sample size of 1015. Females were 70.0% in the source population and 74.7% in the study sample.

The proportion of participants who agreed that vaccines are important for children and for self was 97.4% (95% confidence interval [CI] 96.1% to 98.3%) and 95.4% (95% CI 93.9 to 96.6) respectively. In addition, 95.4% (95% CI 93.8 to 96.6) agreed that vaccines are safe, 97.4% (95% CI 96.15 to 98.28) that vaccines are effective, and 96.1% (95% CI 94.6 to 97.2) that vaccines are compatible with their religious beliefs. The proportion of participants who were willing to receive a COVID-19 vaccine was 89.5% (95% CI 87.2 to 91.4).

Log binomial regression revealed statistically significant positive associations between the intention to receive a COVID-19 vaccine and the belief that vaccines are safe (relative risk [RR] =32.2, CI 4.67 to 221.89); effective (RR=21.4, CI 3.16 to 145.82); important for children (RR=3.5, CI 1.78 to 6.99); important for self (RR=18.5, CI 4.78 to 71.12) or compatible with their religious beliefs (RR=2.2, CI 1.46 to 3.78).

**Conclusion:** Vaccine confidence levels of the study respondents were highly positive. Nevertheless, this could be further enhanced by targeted interventions.

**SUMMARY BOX:** *What is already known?:*

➢ The fast-tracked development and roll out of COVID-19 vaccines has increased general concerns about vaccines
➢ Healthcare workers are critical to the success of any vaccination endeavor.

*What are the new findings?:*

➢ Vaccine sentiments in the study population of healthcare staff and students of Stellenbosch University are highly positive.
➢ The study population has minimal variation in vaccine sentiments and vaccination intention across a range of demographic and social variables.
➢ Log binomial regression identified positive sentiments for all five vaccine sentiments investigated as likely predictors of intention to receive a COVID-19 vaccine when one becomes available.

*What do the new findings imply?:*

➢ Similar interventions can be applied to enhance vaccine confidence among the healthcare staff and students of the study population as both groups share comparable characteristics across socio-demographic spheres.
➢ Strengthening confidence in vaccine importance, safety and effectiveness among healthcare workers and students holds the promise of a potential positive impact on COVID-19 vaccination uptake in the general population in the near and far future.

## INTRODUCTION

### Background

About twenty months since the infamous appearance of the coronavirus disease 2019 (COVID-19) pandemic caused by severe acute respiratory syndrome coronavirus 2 (SARS-CoV-2)[1] on the global stage, the world is still struggling to combat and curtail the disease and its effects. More than 4 million lives of the over 187,419,838 infected by the virus have been lost across the globe as at mid-2021[2]. The emergence of different, more virulent strains of high infectivity of which there are at least four variants currently circulating in different parts of the world[3–6], coupled with multiple waves of the pandemic[7–10], further compound the problem. It has been suggested that to effectively contain the menace of the pandemic, population wide vaccination is crucial[11–13]. It is widely acknowledged that the success of vaccination endeavor depends largely on the healthcare workforce, who usually are the most trusted source of health information for the public[14–16].These healthcare workers were the first to receive a COVID-19 vaccine when one became available [17], and are not immune to vaccination concerns as previously documented[14–16, 18, 19]. The accelerated development, production and pre-licensure emergency use authorization of the successful vaccine candidates has added pressure to the already strained public confidence in vaccines[11, 20]. These may also have inadvertently fueled the vaccination misinformation on web-based information and social media platforms[21–26]. It is against this backdrop of heightened vaccine confidence deficit that healthcare workers are expected to receive, promote and administer successful COVID-19 vaccine candidates to a pandemic-stressed, vaccine wary public.

Moreover, healthcare workers do not operate in a vacuum, they are usually nested within healthcare systems and communities. The resilience of many healthcare systems have been severely tested since the outbreak of the pandemic[27], and the fragile ones, especially those in Africa, have struggled in the current crises[27]. This engenders a situation in which concerned healthcare workers operating within beleaguered healthcare systems have the responsibility of administering and promoting COVID-19 vaccines and vaccination to the general public. It is, however, notable that a significant measure of success has been achieved as millions of doses of the vaccines have been administered on healthcare workers and the general public alike in spite of these challenges.

These issues of concern and challenges highlighted above amongst other reasons previously identified[20], informed the investigation of vaccine confidence levels in a cohort of healthcare workers and their trainers ahead of the COVID-19 vaccination rollout in South Africa.

## METHODS

### Study design

This is a cross sectional survey targeting all academic staff and students of the Faculty of Medicine and Health Sciences at Stellenbosch University in Cape Town, South Africa. Academic staff, defined as staff that are engaged in research or teaching at undergraduate and/or postgraduate levels were the original target of the survey as indicated in the published protocol[20], however, a protocol deviation led to the inclusion of non-teaching, administrative staff.

Participants were informed of their rights to participate or otherwise in the survey invite email, voluntary participation was deemed as implied consent.

### Study population description and sampling strategy

The target population included 4,304 students of which 3,016 (70.1%) are females and 1,287 (30%) males, 1 (0.02%) individual identified as non-binary. This student population consisted of 1599 (37.1%) postgraduates, 2,650 (61.6%) undergraduates and 55(1.3%) occasional students. There were 1,278 staff members in the target population of which 893 (70.0%) were females and 385 (30.0%) were males. These numbers were obtained from the institutions’ information holders.

No sampling strategy was applied as a census of the entire source population was intended. Nevertheless, the promise of a short survey completion time (the survey took an average of 3 minutes) and an incentive of 3 randomly selected participants with complete survey responses winning 500 South African Rands (approximately 25.60 GBP) cash prize were aimed at maximizing the response rate.

### Sample size and response rate

The estimated sample size based on a finite population of between 3,500 and 5,000 potential participants was calculated to be between 1,009 and 1,105; the assumptions underpinning this calculation have been described previously[20].

### Data collection

The survey was conducted online using the Checkbox^®^ survey software on the Stellenbosch University Survey platform to capture participants’ responses. Data collection was conducted between the 4^th^ of February and the 7^th^ of March 2021, prior to the implementation of the COVID-19 vaccine rollout. The tool of data collection was an online administered succinct questionnaire consisting of 6 demographic questions, 5 vaccine confidence statements and 1 intention to receive a COVID-19 vaccine when one becomes available statement. The demographic information elicited included age, sex, religion, academic status (i.e. staff, student or both); highest degree obtained and number of years of schooling post-high school. Four of the 6 statements used to explore vaccine sentiments were obtained directly from the previously validated vaccine confidence statements used in a study conducted in 67 countries[28, 29]. The 4 statements are: “vaccines are important for children to have”; “overall I think vaccines are safe”; “overall I think vaccines are effective”; and “vaccines are compatible with my religious beliefs”. The 2 newly added statements are: “vaccines are important for me to have”, and “I will take a COVID-19 vaccine when one becomes available”. The former explores the individual’s sentiments about the importance of vaccination for oneself; while the latter explores the individuals’ intention to vaccinate with particular reference to the [then] imminent COVID-19 vaccination roll-out. The process of development and the rationale for the inclusion of the two additional questions, has also been previously described[20].

The full questionnaire was pilot tested on a small sample of respondents who shared similar characteristics with the study population but were not included in the study. This was to explore the face validity, feasibility and the logistics of administering the survey. The survey was deemed satisfactory in these three aspects. Each study participant was asked to rate the degree with which he or she agrees with the 6 vaccine confidence statements on a 5-point Likert scale consisting of the following ordinal categories: strongly agree, tend to agree, do not know, tend to disagree, strongly disagree. The questionnaire was in English.

### Data analysis

The complete responses were exported from the data collection platform Checkbox^®^ survey via Stellenbosch University surveys to a Microsoft Excel spreadsheet where initial data cleaning and coding was done. The data set was subsequently exported into IBM^®^ SPSS Statistics software version 27 (IBM Corp. Released 2020. IBM SPSS Statistics for Windows, Version 27.0. Armonk, NY: IBM Corp), where further cleaning, labelling and analysis were performed. EpiCalc 2000 version 1.02 (Joe Gilman and Mark Myatt, Brixton Books, 1998) was used in the calculation of 95% confidence intervals, while Stata/MP version 17.0 for Windows (Stata Corp LLC, Texas, USA) software was used to compute the crude relative risks using log binomial models.

The main outcome investigated is the variability in the vaccine sentiments and vaccine intention within and across all groups investigated. This was done by considering the fraction of respondents that either agree or disagree with the five statements on immunization and the one statement on intention to receive a COVID-19 vaccine. The “strongly agree” and “tend to agree” responses were combined to make up the positive vaccine sentiment variable, while the “strongly disagree” and “tend to disagree” responses were combined to make up the negative vaccine sentiment variable. The “don’t know” responses were removed from the data prior to analyses according to the study’s methodology previously described[20].

Categorical variables were summarized using frequencies and proportions. Quantitative variables are presented using medians and inter quartile ranges as they were not normally distributed. Statistical significance was defined at a p-value <0.05.

Pearson’s chi square exact 2-sided p-values were used to assess the association between categorical variables, while the Mann-Whitney U test was used for the quantitative variables.

The overall vaccine acceptance was >10%, therefore, a log binomial regression model was used to assess the association between vaccine confidence and the intention to receive a COVID-19 vaccine when one becomes available as previously indicated[20]. The strength of this association was assessed using the crude relative risk (RR), and 95% confidence interval (CI) was calculated for each vaccine confidence statement and the intention to receive a COVID-19 vaccine. Due to high levels of multicollinearity, a multivariable model was not possible.

## RESULTS

The total number of estimated potential respondents based on the number of email invites sent out was 4,659 (3,737 students and 922 staff). A total of 1,414 participants responded to the survey invite giving a total response rate of 30.35%. Of the 1,414 responses received, 1,015 were complete responses. This gives an actual response rate of 21.79%.

The age of the 1015 respondents that submitted complete survey responses (study sample population) ranged from 16 to 90 years with a median of 53 years. An overview of the characteristics of the study population is provided in Table 1.

**Table 1:** Characteristics of the study sample population

| Demographic variables |  | Count | Percentage |
| --- | --- | --- | --- |
| Staff/Student | Staff | 257 | 25.3 |
|  | Student | 675 | 66.5 |
|  | Both | 83 | 8.2 |
| Sex | Male | 255 | 25.1 |
|  | Female | 758 | 74.7 |
|  | Other | 2 | 0.02 |
| Degree | BSc | 370 | 36.5 |
|  | Hons | 135 | 13.3 |
|  | MBBS | 219 | 21.6 |
|  | MSc | 200 | 19.7 |
|  | PhD | 91 | 9.0 |
| Religion | Islam | 111 | 11.0 |
|  | Roman Catholic | 91 | 9.0 |
|  | Orthodox | 23 | 2.2 |
|  | Pentecostal | 155 | 15.0 |
|  | Traditional | 75 | 7.4 |
|  | Jewish | 8 | 0.8 |
|  | Buddhist | 4 | 0.4 |
|  | Hindu | 25 | 2.5 |
|  | Atheist | 76 | 7.5 |
|  | Agnostic | 73 | 7.2 |
|  | Other | 365 | 36.0 |
|  | 7th Day Adventist | 9 | 1.0 |
| Post matriculation years of schooling | $\leq 5$ | 449 | 44.2 |
|  | 6-10 | 283 | 27.9 |
|  | 11-14 | 147 | 14.5 |
|  | 15-20 | 52 | 5.1 |
| | $\geq 21$ | 84 | 8.3 |
| Age group | $\leq 24$ | 414 | 40.8 |
|  | 25-34 | 243 | 23.9 |
|  | 35-44 | 193 | 19.0 |
|  | 45-54 | 84 | 8.3 |
|  | 55-64 | 70 | 6.9 |
| | $\geq 65$ | 11 | 1.1 |

*The total number of valid responses was 1015 for all categories. Values given are absolute counts and percentages for each variable.*

The total complete responses for the five vaccine confidence statements and the intention to receive a vaccine COVID-19 vaccine are shown in Table 2. A strong internal consistency exists within the 5 vaccine confidence statements as revealed by the Cronbach’s alpha of 0.840.

**Table 2:** Vaccine sentiments among the study population

| **Statements** |  |  |  |  |  |
| --- | --- | --- | --- | --- | --- |
|  | Strongly agree | Tend to agree | Don't know | Tend to disagree | Strongly disagree |
| 1. Vaccines are important for children to have | 862<br>(84.93%) | 107<br>(10.54%) | 20<br>(1.97%) | 19<br>(1.87%) | 7<br>(0.69%) |
| 2. Vaccines are important for me to have | 760<br>(74.88%) | 180<br>(17.73%) | 30<br>(2.96%) | 32<br>(3.15%) | 13<br>(1.28%) |
| 3. Overall, I think vaccines are safe | 560<br>(55.17%) | 335<br>(33.00%) | 77<br>(7.59%) | 31<br>(3.05%) | 12<br>(1.18%) |
| 4. Overall, I think vaccines are effective | 603<br>(59.41%) | 338<br>(33.30%) | 49<br>(4.83%) | 13<br>(1.28%) | 12<br>(1.18%) |
| 5. Vaccines are compatible with my religious beliefs | 704<br>(69.36%) | 180<br>(17.73%) | 95<br>(9.36%) | 19<br>(1.87%) | 17<br>(1.67%) |
| 6. I will take a Covid-19 vaccine when one becomes available | 631<br>(62.17%) | 150<br>(14.78%) | 142<br>(13.99%) | 46<br>(4.53%) | 46<br>(4.53%) |

*The total number of valid responses was 1015 for all statements. Values given are absolute counts (percentages) for each response*

Overall, the study participants showed high level of agreement with the five vaccine confidence statements. There was good precision in the estimates, of ± 3%. However, the percentage of people who intend to receive a COVID-19 vaccine was slightly lower at 89.5% (95% CI 87.19 to 91.38).

The results of the associations between vaccine confidence and intention are shown in Table 4. Participants who expressed positive vaccine sentiments for vaccine safety were 32 times (95% CI 4.67 to 221.89) more likely to agree to receive a COVID-19 vaccine. A similar pattern was observed for the other vaccine sentiments. However, the compatibility of vaccine with religion statement showed that the likelihood of those participants who agree that vaccines are compatible with their religious belief to indicate their willingness to receive a COVID-19 vaccine to be 2.2. Nevertheless, the effect size had a comparatively higher precision at 95% CI of 1.46 to 3.78. This trend was also observed with the “vaccines are important for children to have” statement which had a slightly higher relative risk of 3.5 (95% CI 1.78 to 6.99). The p value for the association between each of the five vaccine confidence statements and the intention to receive a COVID-19 vaccine was p<0.001 as shown in Table 4.

**Table 3:** The percentage of agreement with the vaccine confidence statements and intention to receive a COVID-19 vaccine

| **Statements** | **Agree** | **Confidence interval**<br>**for agreement** |
| --- | --- | --- |
| 1. Vaccines are important for children to have | 97.4%<br>(n=969) | 96.14 – 98.25 |
| 2. Vaccines are important for me to have | 95.4%<br>(n=940) | 93.88 – 96.61 |
| 3. Overall, I think vaccines are safe | 95.4%<br>(n= 895) | 93.82 – 96.62 |
| 4. Overall, I think vaccines are effective | 97.4%<br>(n=941) | 96.15 – 98.28 |
| 5. Vaccines are compatible with my religious beliefs | 96.1%<br>(n=884) | 94.57 – 97.21 |
| 6. I will take a Covid-19 vaccine when one becomes available | 89.5%<br>(n=781) | 87.19 - 91.38 |

**Table 4:** Association between vaccine confidence statements and intention to receive a COVID-19 vaccine

| Vaccine confidence statements | Vaccine sentiments | Agree | Disagree | Total | Crude relative risk (95% CI) | P-value |
| --- | --- | --- | --- | --- | --- | --- |
| 1. Vaccines are important for children to have | Agree | 91.7% (n=767) | 8.3% (n=69) | 100% (n=836) | 3.5 (1.78-6 .99) | <0.001 |
|  | Disagree | 26.1% (n=6) | 73.9% (n=17) | 100% (n=23) |  |  |
|  | Total | 90% (n= 773) | 10% (n=86) | 100% (n=859) |  |  |
| 2. Vaccines are important for me to have | Agree | 94.6% (n=772) | 5.4% (n=44) | 100% (n=816) | 18.5 (4.78-71.12) | <0.001 |
|  | Disagree | 5.1% (n=2) | 94.9% (n=37) | 100% (n=39) |  |  |
|  | Total | 90.5% (n=774) | 9.5% (n=81) | 100% (n=855) |  |  |
| 3. Overall, I think vaccines are safe | Agree | 94.7% (n=749) | 5.3% (n=42) | 100% (n=791) | 32.2 (4.67-221 .89) | <0.001 |
|  | Disagree | 2.9% (n=1) | 97.1% (n=33) | 100% (n=34) |  |  |
|  | Total | 90.9% (n=750) | 9.1% (n=75) | 100% (n=825) |  |  |
| 4. Overall, I think vaccines are effective | Agree | 93.3% (n=762) | 6.7% (n=55) | 100% (n=817) | 21.4 (3.16-145 .82) | <0.001 |
|  | Disagree | 4.3% (n=1) | 95.7% (n=22) | 100% (n=23) |  |  |
|  | Total | 90.8% (n=763) | 9.2% (n=77) | 100% (n=840) |  |  |
| 5. Vaccines are compatible with my religious beliefs | Agree | 93.2% (n=715) | 6.8% (n=52) | 100% (n=767) | 2.2 (1.46-3.78) | <0.001 |
|  | Disagree | 41.4% (n=12) | 58.6% (n=17) | 100% (n=29) |  |  |
|  | Total | 91.3% (n=727) | 8.7% (n=69) | 100% (n=796) |  |  |

For categorical demographic variables, no potential predictors were found associated with either the vaccine confidence statements or the intention to receive a COVID-19 vaccine statement at the bivariate level of analysis, as shown in Tables 5-13 (supplementary files).

The distributions of quantitative variables between those who agreed with and those who disagreed with the vaccine confidence statements and with the intention to receive a COVID-19 vaccine when one becomes available were not different between the groups as shown in the supplementary files. Furthermore, there was no significant statistical association between any quantitative variables and vaccine confidence.

Overall, and within all demographic categories, the vaccine sentiment of the study respondents was highly positive. Most categories of respondents reported over 90% agreements with the 5 vaccine confidence statements. The exception (a lower than 90% level of agreement) was the 7^th^ Day Adventist group in the religion category (7 individuals) which had 66.7% agreement with vaccine confidence statements 2 and 3 (‘vaccines are important for me to have’ and ‘overall, I think vaccines are safe’), this is shown in tables 7 and 9 in the supplementary files. They also had a 77.8% agreement with vaccine confidence statement 4 (overall, I think vaccines are effective) as reflected in table 11 in the supplementary files. However, this same category of religion had 100% agreement that vaccines are compatible with their religious beliefs (vaccine confidence statement 5) as shown in table 13 in the supplementary files, and that vaccines are important for children to have (vaccine confidence statement 1) as reflected in table 5 in the supplementary files. Any trends for this group should be interpreted with caution because of the small numbers involved.

Positive vaccine sentiments were comparably slightly less across all categorical variables for the intention to receive a COVID-19 vaccine when one becomes available than it was for the five vaccine confidence statements. Table 15 shows this with many of the factors having >80%, but mostly less than 90% agreement with the COVID-19 vaccine receipt intention. Again, the exception was the 7^th^ Day Adventist group (only 7 persons) which had only 42.9% agreement with the statement for the receipt of a COVID-19 vaccine when one becomes available. This result is shown in table 15 in the supplementary files.

Due to the low numbers per cells and the many groups within the religion category, the p values for vaccine confidence statements 2 and 3, and the intention to receive a COVID-19 vaccine when one becomes available statement could not be computed. This can be seen in tables 7, 9 and 15 in the supplementary files. Nevertheless, there is negligible evidence to suggest that if these p values had been computed, they would have been different from the other p values obtained for the religion category for the other vaccine confidence statements. An example of this is shown in table 13, where the p-value for the religion category is 0.851.

No significant association between levels of education and vaccine statements were detected either at the postgraduate (Honors & above) or undergraduate (BSc. & MBBS) level. Moreover, the level of positive vaccine sentiments expressed by both levels of education is very high, in all cases >90% as shown in table 17a in the supplementary files. This table also shows that individuals with either level of education were equally persuaded that vaccines are compatible with their religious beliefs (p=1.000). The positive sentiment expressed for the intention to receive a COVID-19 vaccine when one becomes available was slightly lower than those expressed for the vaccine confidence statements, it was nevertheless, high at >89%. No association was found between levels of education and the intention to receive a COVID-19 vaccine when one becomes available (p=1.000).

The re-categorization of levels of education to Honors degree or lower versus Masters and above led to a significant association between the level of education and the belief that vaccines are important for the respondent to have (p=0.043). A slightly lower percentage of those with up to Honors education agreed that vaccines were important for them to have, as compared with those with Master’s degree and higher (94.5% vs 97.6%), however this small difference might not be practically important. A high level of positive vaccine sentiments was expressed by both levels of education with a >90% agreement with the all vaccine confidence statements. However, a slightly lesser proportion of the up to Honors group (88.4%) indicated their willingness to take a COVID-19 vaccine when one becomes available compared to the 92.0% of the Masters and above group but this difference was not statistically significant.

## DISCUSSION

In this study, the vaccine confidence levels of healthcare workers in training and staff of the named training institution were estimated.

Our results show high levels of positive vaccine sentiments expressed by the study respondents across and within all groups investigated. This implies high levels of confidence in the vaccine constructs investigated which are: importance for children and self, safety, effectiveness, and compatibility with religious beliefs. The intention to receive a COVID-19 vaccine when one becomes available was also investigated. Although vaccination intention does not always translate to action[30, 31], nevertheless, the positive disposition of current and future healthcare workers of the study population is potentially indicative of better promotion of uptake of routine and COVID-19 vaccines and vaccination. This is assumed because of the important influence of healthcare workers as a credible source of information to the public on routine childhood vaccines[14–16] and more recently COVID-19 vaccines[32]. Our results are also consistent with the findings of a similar study that reported on vaccine sentiments of the more educated, urban dwelling online adult population portion of the general South African public[33]. While this 2020 survey reported that 64% of their described study population are willing to receive a COVID-19 vaccine when one becomes available, our results reports a higher 89.5%. This could be due to the relatively homogenous nature of our study population, and the more recent timing of our study. The findings of our study is congruent with that of Lazarus *et al*[31] that reported that 81.58% of the South African population included in their study indicated their willingness to receive a COVID-19 vaccine; and further supported by the results of another recent study, which found that an estimated 79% of Africans are willing to receive a COVID-19 vaccine[34]. Results from surveys conducted in other parts of the world also report high levels of willingness to receive a COVID-19 vaccine[31, 32, 35, 36].

The high level of intention to receive a COVID-19 vaccine reported in our study was however comparatively less than those reported for the 5 vaccine confidence statements, all of which were ≥95%. One reason we are suggesting for this is that COVID-19, being a relatively new disease, does not yet have its medium and long term prognosis well understood and defined like other vaccine preventable diseases (VPDs) for which vaccines are routinely administered and that the vaccines are newly developed. However, this may not be the only reason for this observed trend.

A log binomial regression of the intention to receive a COVID-19 vaccine when one becomes available (the outcome of interest) against the 5 vaccine statements revealed confidence in the safety and effectiveness of such vaccines as the most likely predictors of the intention to receive it. This finding is noteworthy as safety and to a lesser degree, effectiveness concerns has been cited in literature as some of the leading reasons for reluctance to receive a COVID-19 vaccine[30, 32, 37, 38]. In seeming contradiction, the consideration of safety and effectiveness of a COVID-19 vaccine has also been proffered as reasons to receive the vaccines[31, 39–42]. These two constructs are pivotal to the acceptance or otherwise of any vaccine, and are often included in the items used to query the intention to receive a COVID-19 vaccine as exemplified by the study of Lazarus *et al*[31] and the Africa Centre for Disease Control and Prevention (Africa CDC) survey[34].

With particular regards to COVID-19 vaccines, caregivers were reported to be more willing to vaccinate their charges than themselves[39], while the opposite was reported about healthcare workers as parents[43]. Therefore, the high levels of positive sentiments expressed by our study participants particularly for the importance of vaccines for self and children could be considered as a potential indication of better vaccine uptake and coverage in the population that will be served by these healthcare workers in the future.

Religious beliefs can have a positive or negative influence on vaccination activities as previously documented in literature[44–48]. In our study population, respondents in the various religious groups investigated showed high levels of positive vaccine sentiments, with the exception of the one highlighted earlier. Respondents who consider vaccines to be compatible with their religious belief were twice as likely to indicate their willingness to receive a COVID-19 vaccine when one becomes available.

There was marginal variation in the level of the other demographic factors, with most showing consistently high levels of agreements with the 5 vaccine confidence statements and a slight decrease in the intention to receive a COVID-19 vaccine as shown in the supplementary files. Staff members showed slightly higher levels of agreements with the 3 out of the 5 vaccine confidence statements and the intention to receive a COVID-19 vaccine than students or the ‘both staff and student’ group, they had equal levels of agreement with the students with vaccine efficacy statement which was minimally higher than that of the ‘both staff and student’ group. Only in the “vaccine is compatible with my religious belief” statement did the student group have a slightly higher agreement level than the staff and ‘both’ groups (96.7% versus (94.8% & 94.9% respectively). This variation in levels of agreement (indicative of levels of confidence) though minimal and not statistically significant, is, nevertheless important. It shows that staff members having higher levels of vaccine confidence are rightly positioned to influence and impact future healthcare workers while they are still in training. This further reinforces the anticipation that future healthcare workers will have positive vaccine sentiments, which in turn, should translate to better vaccine recommendation to and vaccine uptake in the population served by such healthcare workers[16, 37, 49].

The male respondents in our study showed a marginally, but not statistically significant higher levels of agreements with 3 out of the 5 vaccine statements. The exceptions to this trend being the “vaccine are important for children to have” and the “vaccines are compatible with my religious belief statements”. The reasons for the exceptions could be that females are generally the primary caregivers to children (especially infants and pre-teens), and, as anecdotal evidence suggests, females are more inclined to religious activities than males. Nevertheless, this finding aligns with the findings of some recently published studies[43, 50– 52], but is in contrast with the report in the Kaiser Health News[53] which found females more willing than males to receive a COVID-19 vaccine in the United States of America.

The age and number of years of post-high school schooling also follows a similar trend to those of the other demographic factors. The participants who agreed with the 5 vaccine statements and the intention to receive a COVID-19 vaccine when one becomes available had a median of 6.0 number of years of post-high school schooling. In contrast, the median years of those who disagree with the vaccine confidence statements and intention to receive a COVID-19 vaccine oscillated between 5.0 and 5.5. The median age distribution in years for the participants that agree with the 5 vaccine confidence statements was consistently 29, and 30 for those who indicated their willingness to receive a COVID-19 vaccine. On the contrary, the median age for those who disagree varied considerably, from 24 to 33 as shown in the supplementary files. Nonetheless, it was also 30 years for those who indicated their unwillingness to receive a COVID-19 vaccine.

This trend changed when the ages were grouped together. The age group ≤ 24 years showed a lesser degree of agreement with 3 out of the 5 vaccine confidence statements, only in the “vaccines are important for children to have” and “vaccines are compatible with my religious beliefs” did they have a slightly higher levels of agreements than the ≥65 year olds. These, on the other hand, were more likely to indicate their willingness to receive a COVID-19 vaccine when one becomes available. This finding is also supported by recent literature[37, 50, 51, 54].

Our study found no significant association between levels of education (undergraduates and post graduates) and 4 of the 5 vaccine confidence statements. These findings are consistent with previous studies that found higher levels of education of healthcare workers to be associated with greater vaccine confidence[37, 49, 55]. The level of education in our study population can be considered to be relatively high, as most respondents have at least some form of post high school schooling. Therefore, the findings of our study should be viewed with this in mind.

This study is not without limitations. Low response rate characteristic of online surveys is one of such limitations. However, the actual response rate of 21.79% which gave an actual sample size of 1015 was sufficient providing adequate precision for the study as it was within the estimated sample size range calculated in the published protocol[20]. The study population was similar to the target population, using gender distribution as indicator. Nevertheless, it should be noted that the views expressed by the study sample population may not adequately represent the views those not supportive of vaccination in the target population. Thus, the possibility of selection bias cannot be ruled out due to the voluntary nature of the survey. No diploma and not all possible degrees were included in the response options for the question “highest degree obtained” and this may have some influence on the response to the question.

## CONCLUSION

We conclude that the levels of vaccine confidence in our study population were very high in the immediate period prior to the initiation of the vaccine programme in South Africa. Nevertheless, more emphasis on vaccine education and promotion will be beneficial among these current and future healthcare workers. This will potentially further enhance vaccine acceptance and uptake in the general population in the short, medium and long term.

## Data Availability

All data relevant to the study are included in the article or uploaded as supplementary information.

## ETHICS APPROVAL

This study obtained ethics approval from Stellenbosch University in South Africa: Human Research Ethics Committee (HREC) reference # S19/01/014 (PhD). Participation in the survey is deemed as implied consent.

### Author contributions

EO led the conceptualization, design, and drafted the manuscript. TE conducted the statistical analysis of the data and gave feedback on the manuscript; HM and CSW provided the supervisory overview and feedback on the overall study, methodology, and critically reviewed the manuscript. This team of four authors gives their approval for the publishing of this original research manuscript.

## Acknowledgements

The corresponding author, E.O. acknowledges the efforts of Dr Edward Nicol, Dr Samuel Egieyeh and Ms. Naomi Okugbeni-Doghor who assisted in different ways in the course of the study.

## Patient and public involvement

Patients and the public were not involved in the design, execution, and analysis of the study.

## Patient consent

Not required.

## Provenance and peer review

Not commissioned; externally peer reviewed.

## Open access

This is an open access article distributed in accordance with the Creative Commons Attribution Non Commercial (CC BY-NC 4.0) license, which permits others to distribute, remix, adapt, build upon this work non-commercially, and license their derivative works on different terms, provided the original work is properly cited, appropriate credit is given, any changes made indicated, and the use is non-commercial. See: http://creativecommons.org/licenses/by-nc/4.0/.

