## Supplementry Tables for "Estimating vaccine confidence levels among healthcare staff and students of a tertiary institution in South Africa"

*Table 5:* Associations between categorical demographic variables and the importance of vaccines for children statement

| Categorical demographic variables | | Vaccines are important for children to have statement | | | | | | p-value |
| --- | --- | --- | --- | --- | --- | --- | --- | --- |
|  |  | Disagree | | Agree | | Total | |  |
|  |  | Count | Row N % | Count | Row N % | Count | Row N % |  |
| Staff/Student | Staff | 4 | 1.6% | 245 | 98.4% | 249 | 100.0% | 0.472 |
|  | Student | 19 | 2.9% | 644 | 97.1% | 663 | 100.0% |  |
|  | Both | 3 | 3.6% | 80 | 96.4% | 83 | 100.0% |  |
|  | Total | 26 | 2.6% | 969 | 97.4% | 995 | 100.0% |  |
| Sex | Male | 9 | 3.6% | 241 | 96.4% | 250 | 100.0% | 0.376 |
|  | Female | 17 | 2.3% | 727 | 97.7% | 744 | 100.0% |  |
|  | Other | 0 | 0.0% | 1 | 100.0% | 1 | 100.0% |  |
|  | Total | 26 | 2.6% | 969 | 97.4% | 995 | 100.0% |  |
| degree | BSc | 10 | 2.8% | 345 | 97.2% | 355 | 100.0% | 0.786 |
|  | Hons | 5 | 3.8% | 128 | 96.2% | 133 | 100.0% |  |
|  | MBBS | 6 | 2.8% | 212 | 97.2% | 218 | 100.0% |  |
|  | MSc | 3 | 1.5% | 196 | 98.5% | 199 | 100.0% |  |
|  | PhD | 2 | 2.2% | 88 | 97.8% | 90 | 100.0% |  |
|  | Total | 26 | 2.6% | 969 | 97.4% | 995 | 100.0% |  |
| religion | Islam | 7 | 6.4% | 103 | 93.6% | 110 | 100.0% | 0.351 |
|  | Roman Catholic | 3 | 3.3% | 88 | 96.7% | 91 | 100.0% |  |
|  | Orthodox | 10 | 3.2% | 303 | 96.8% | 313 | 100.0% |  |
|  | Pentecostal | 3 | 1.6% | 186 | 98.4% | 189 | 100.0% |  |
|  | Traditional | 2 | 2.7% | 73 | 97.3% | 75 | 100.0% |  |
|  | Jewish | 0 | 0.0% | 7 | 100.0% | 7 | 100.0% |  |
|  | Buddhist | 0 | 0.0% | 4 | 100.0% | 4 | 100.0% |  |
|  | Hindu | 0 | 0.0% | 25 | 100.0% | 25 | 100.0% |  |
|  | Atheist | 0 | 0.0% | 74 | 100.0% | 74 | 100.0% |  |
|  | Agnostic | 1 | 1.4% | 72 | 98.6% | 73 | 100.0% |  |
|  | Other | 0 | 0.0% | 25 | 100.0% | 25 | 100.0% |  |
|  | 7th Day Adventist | 0 | 0.0% | 9 | 100.0% | 9 | 100.0% |  |
|  | Total | 26 | 2.6% | 969 | 97.4% | 995 | 100.0% |  |
| Age group | ≤24 | 13 | 3.2% | 394 | 96.8% | 407 | 100.0% | 0.457 |
|  | 25-34 | 3 | 1.2% | 238 | 98.8% | 241 | 100.0% |  |
|  | 35-44 | 5 | 2.7% | 183 | 97.3% | 188 | 100.0% |  |
|  | 45-54 | 2 | 2.5% | 78 | 97.5% | 80 | 100.0% |  |
|  | 55-64 | 2 | 2.9% | 67 | 97.1% | 69 | 100.0% |  |
|  | ≥65 | 1 | 10.0% | 9 | 90.0% | 10 | 100.0% |  |
|  | Total | 26 | 2.6% | 969 | 97.4% | 995 | 100.0% |  |

*Table 6:* Associations between quantitative variables and importance of vaccines for children statement

| Quantitative variables | | Vaccines are important for children to have statement | | |  |
| --- | --- | --- | --- | --- | --- |
|  |  | Disagree | Agree | Total | p-value |
| age | Median | 24.00 | 29.00 | 29.00 | 0.523 |
|  | Percentile 25 | 20.00 | 22.00 | 22.00 |  |
|  | Percentile 75 | 39.00 | 38.00 | 38.00 |  |
| Post matric years of schooling | Median | 5.50 | 6.00 | 6.00 | 0.572 |
|  | Percentile 25 | 3.00 | 4.00 | 4.00 |  |
|  | Percentile 75 | 13.00 | 11.00 | 11.00 |  |

*Table 7:* Associations between categorical demographic variables and importance of vaccines for self statement

| Categorical demographic variables | | Vaccines are important for me to have statement | | | | | | p-value |
| --- | --- | --- | --- | --- | --- | --- | --- | --- |
|  |  | Disagree | | Agree | | Total | |  |
|  |  | Count | Row N % | Count | Row N % | Count | Row N % |  |
| Staff/Student | Staff | 10 | 4.0% | 238 | 96.0% | 248 | 100.0% | 0.882 |
|  | Student | 31 | 4.7% | 626 | 95.3% | 657 | 100.0% |  |
|  | Both | 4 | 5.0% | 76 | 95.0% | 80 | 100.0% |  |
|  | Total | 45 | 4.6% | 940 | 95.4% | 985 | 100.0% |  |
| Sex | Male | 9 | 3.6% | 240 | 96.4% | 249 | 100.0% | 0.485 |
|  | Female | 36 | 4.9% | 700 | 95.1% | 736 | 100.0% |  |
|  | Total | 45 | 4.6% | 940 | 95.4% | 985 | 100.0% |  |
| degree | BSc | 19 | 5.4% | 334 | 94.6% | 353 | 100.0% | 0.140 |
|  | Hons | 10 | 7.8% | 119 | 92.2% | 129 | 100.0% |  |
|  | MBBS | 9 | 4.2% | 206 | 95.8% | 215 | 100.0% |  |
|  | MSc | 4 | 2.0% | 194 | 98.0% | 198 | 100.0% |  |
|  | PhD | 3 | 3.3% | 87 | 96.7% | 90 | 100.0% |  |
|  | Total | 45 | 4.6% | 940 | 95.4% | 985 | 100.0% |  |
| religion | Islam | 7 | 6.5% | 101 | 93.5% | 108 | 100.0% |  |
|  | Roman Catholic | 4 | 4.3% | 88 | 95.7% | 92 | 100.0% |  |
|  | Orthodox | 12 | 3.9% | 295 | 96.1% | 307 | 100.0% |  |
|  | Pentecostal | 10 | 5.3% | 179 | 94.7% | 189 | 100.0% |  |
|  | Traditional | 4 | 5.6% | 68 | 94.4% | 72 | 100.0% |  |
|  | Jewish | 0 | 0.0% | 7 | 100.0% | 7 | 100.0% |  |
|  | Buddhist | 0 | 0.0% | 4 | 100.0% | 4 | 100.0% |  |
|  | Hindu | 0 | 0.0% | 25 | 100.0% | 25 | 100.0% |  |
|  | Atheist | 2 | 2.6% | 74 | 97.4% | 76 | 100.0% |  |
|  | Agnostic | 2 | 2.8% | 70 | 97.2% | 72 | 100.0% |  |
|  | Other | 1 | 4.2% | 23 | 95.8% | 24 | 100.0% |  |
|  | 7th Day Adventist | 3 | 33.3% | 6 | 66.7% | 9 | 100.0% |  |
|  | Total | 45 | 4.6% | 940 | 95.4% | 985 | 100.0% |  |
| Age group | ≤24 | 18 | 4.5% | 386 | 95.5% | 404 | 100.0% | 0.463 |
|  | 25-34 | 8 | 3.4% | 225 | 96.6% | 233 | 100.0% |  |
|  | 35-44 | 10 | 5.2% | 181 | 94.8% | 191 | 100.0% |  |
|  | 45-54 | 3 | 3.8% | 77 | 96.3% | 80 | 100.0% |  |
|  | 55-64 | 6 | 9.0% | 61 | 91.0% | 67 | 100.0% |  |
|  | ≥65 | 0 | 0.0% | 10 | 100.0% | 10 | 100.0% |  |
|  | Total | 45 | 4.6% | 940 | 95.4% | 985 | 100.0% |  |

*Table 8:* Associations between quantitative variables and importance of vaccines for self statement

| Quantitative variables | | Vaccines are important for me to have statement | | | p-value |
| --- | --- | --- | --- | --- | --- |
|  |  | Disagree | Agree | Total |  |
| age | Median | 32,00 | 29,00 | 29,00 | 0.429 |
|  | Percentile 25 | 22,00 | 21,00 | 22,00 |  |
|  | Percentile 75 | 39,00 | 38,00 | 38,00 |  |
| Post matric | Median | 5,00 | 6,00 | 6,00 | 0.611 |
|  | Percentile 25 | 3,00 | 4,00 | 4,00 |  |
|  | Percentile 75 | 14,00 | 11,00 | 11,00 |  |

*Table 9:* Associations between categorical demographic variables and vaccine safety statement

| Categorical demographic variables | | Overall, I think vaccines are safe | | | | | | p-value |
| --- | --- | --- | --- | --- | --- | --- | --- | --- |
|  |  | Disagree | | Agree | | Total | |  |
|  |  | Count | Row N % | Count | Row N % | Count | Row N % |  |
| Staff/Student | Staff | 7 | 3.0% | 227 | 97.0% | 234 | 100.0% | 0.379 |
|  | Student | 31 | 5.0% | 592 | 95.0% | 623 | 100.0% |  |
|  | Both | 5 | 6.2% | 76 | 93.8% | 81 | 100.0% |  |
|  | Total | 43 | 4.6% | 895 | 95.4% | 938 | 100.0% |  |
| Sex | Male | 6 | 2.5% | 234 | 97.5% | 240 | 100.0% | 0.118 |
|  | Female | 37 | 5.3% | 660 | 94.7% | 697 | 100.0% |  |
|  | Other | 0 | 0.0% | 1 | 100.0% | 1 | 100.0% |  |
|  | Total | 43 | 4.6% | 895 | 95.4% | 938 | 100.0% |  |
| degree | BSc | 15 | 4.6% | 311 | 95.4% | 326 | 100.0% | 0.285 |
|  | Hons | 10 | 8.3% | 110 | 91.7% | 120 | 100.0% |  |
|  | MBBS | 7 | 3.3% | 207 | 96.7% | 214 | 100.0% |  |
|  | MSc | 7 | 3.7% | 181 | 96.3% | 188 | 100.0% |  |
|  | PhD | 4 | 4.4% | 86 | 95.6% | 90 | 100.0% |  |
|  | Total | 43 | 4.6% | 895 | 95.4% | 938 | 100.0% |  |
| religion | Islam | 7 | 6.8% | 96 | 93.2% | 103 | 100.0% |  |
|  | Roman Catholic | 5 | 5.7% | 82 | 94.3% | 87 | 100.0% |  |
|  | Orthodox | 12 | 4.2% | 277 | 95.8% | 289 | 100.0% |  |
|  | Pentecostal | 9 | 5.1% | 169 | 94.9% | 178 | 100.0% |  |
|  | Traditional | 3 | 4.3% | 66 | 95.7% | 69 | 100.0% |  |
|  | Jewish | 0 | 0.0% | 8 | 100.0% | 8 | 100.0% |  |
|  | Buddhist | 0 | 0.0% | 4 | 100.0% | 4 | 100.0% |  |
|  | Hindu | 0 | 0.0% | 25 | 100.0% | 25 | 100.0% |  |
|  | Atheist | 2 | 2.7% | 72 | 97.3% | 74 | 100.0% |  |
|  | Agnostic | 1 | 1.4% | 69 | 98.6% | 70 | 100.0% |  |
|  | Other | 1 | 4.5% | 21 | 95.5% | 22 | 100.0% |  |
|  | 7th Day Adventist | 3 | 33.3% | 6 | 66.7% | 9 | 100.0% |  |
|  | Total | 43 | 4.6% | 895 | 95.4% | 938 | 100.0% |  |
| Age group | ≤24 | 19 | 5.0% | 364 | 95.0% | 383 | 100.0% | 0.746 |
|  | 25-34 | 8 | 3.6% | 217 | 96.4% | 225 | 100.0% |  |
|  | 35-44 | 8 | 4.4% | 172 | 95.6% | 180 | 100.0% |  |
|  | 45-54 | 3 | 4.0% | 72 | 96.0% | 75 | 100.0% |  |
|  | 55-64 | 5 | 7.7% | 60 | 92.3% | 65 | 100.0% |  |
|  | ≥65 | 0 | 0.0% | 10 | 100.0% | 10 | 100.0% |  |
|  | Total | 43 | 4.6% | 895 | 95.4% | 938 | 100.0% |  |

*Table 10:* Associations between quantitative variables and vaccine safety statement

| Quantitative variables | | Overall, I think vaccines are safe | | | p-value |
| --- | --- | --- | --- | --- | --- |
|  |  | Disagree | Agree | Total |  |
| Age | Median | 27,00 | 29,00 | 29,00 | 0.994 |
|  | Percentile 25 | 21,00 | 22,00 | 22,00 |  |
|  | Percentile 75 | 42,00 | 38,00 | 38,00 |  |
| Post matric | Median | 5,00 | 6,00 | 6,00 | 0.323 |
|  | Percentile 25 | 3,00 | 4,00 | 4,00 |  |
|  | Percentile 75 | 13,00 | 11,00 | 11,00 |  |

*Table 11:* Associations between categorical demographic variables and vaccine effectiveness statement

| Categorical demographic variables | | Overall, I think vaccines are effective | | | | | | p-value |
| --- | --- | --- | --- | --- | --- | --- | --- | --- |
|  |  | Disagree | | Agree | | Total | |  |
|  |  | Count | Row N % | Count | Row N % | Count | Row N % |  |
| Staff/Student | Staff | 5 | 2.1% | 235 | 97.9% | 240 | 100.0% | 0.109 |
|  | Student | 15 | 2.3% | 629 | 97.7% | 644 | 100.0% |  |
|  | Both | 5 | 6.1% | 77 | 93.9% | 82 | 100.0% |  |
|  | Total | 25 | 2.6% | 941 | 97.4% | 966 | 100.0% |  |
| Sex | Male | 3 | 1.2% | 243 | 98.8% | 246 | 100.0% | 0.183 |
|  | Female | 22 | 3.1% | 697 | 96.9% | 719 | 100.0% |  |
|  | Other | 0 | 0.0% | 1 | 100.0% | 1 | 100.0% |  |
|  | Total | 25 | 2.6% | 941 | 97.4% | 966 | 100.0% |  |
| degree | BSc | 6 | 1.8% | 333 | 98.2% | 339 | 100.0% | 0.217 |
|  | Hons | 8 | 6.3% | 119 | 93.7% | 127 | 100.0% |  |
|  | MBBS | 5 | 2.3% | 212 | 97.7% | 217 | 100.0% |  |
|  | MSc | 4 | 2.1% | 189 | 97.9% | 193 | 100.0% |  |
|  | PhD | 2 | 2.2% | 88 | 97.8% | 90 | 100.0% |  |
|  | Total | 25 | 2.6% | 941 | 97.4% | 966 | 100.0% |  |
| religion | Islam | 5 | 4.6% | 104 | 95.4% | 109 | 100.0% | 0.109 |
|  | Roman Catholic | 2 | 2.2% | 89 | 97.8% | 91 | 100.0% |  |
|  | Orthodox | 7 | 2.3% | 293 | 97.7% | 300 | 100.0% |  |
|  | Pentecostal | 5 | 2.7% | 180 | 97.3% | 185 | 100.0% |  |
|  | Traditional | 2 | 2.9% | 66 | 97.1% | 68 | 100.0% |  |
|  | Jewish | 0 | 0.0% | 8 | 100.0% | 8 | 100.0% |  |
|  | Buddhist | 0 | 0.0% | 4 | 100.0% | 4 | 100.0% |  |
|  | Hindu | 0 | 0.0% | 25 | 100.0% | 25 | 100.0% |  |
|  | Atheist | 1 | 1.4% | 73 | 98.6% | 74 | 100.0% |  |
|  | Agnostic | 1 | 1.4% | 70 | 98.6% | 71 | 100.0% |  |
|  | Other | 0 | 0.0% | 22 | 100.0% | 22 | 100.0% |  |
|  | 7th Day Adventist | 2 | 22.2% | 7 | 77.8% | 9 | 100.0% |  |
|  | Total | 25 | 2.6% | 941 | 97.4% | 966 | 100.0% |  |
| Age group | ≤24 | 8 | 2.0% | 390 | 98.0% | 398 | 100.0% | 0.486 |
|  | 25-34 | 7 | 3.1% | 221 | 96.9% | 228 | 100.0% |  |
|  | 35-44 | 4 | 2.2% | 180 | 97.8% | 184 | 100.0% |  |
|  | 45-54 | 2 | 2.5% | 78 | 97.5% | 80 | 100.0% |  |
|  | 55-64 | 4 | 6.1% | 62 | 93.9% | 66 | 100.0% |  |
|  | ≥65 | 0 | 0.0% | 10 | 100.0% | 10 | 100.0% |  |
|  | Total | 25 | 2.6% | 941 | 97.4% | 966 | 100.0% |  |

*Table 12:* Associations between quantitative variables and vaccine effectiveness statement

| Quantitative variables | | Overall, I think vaccines are effective | | | p-value |
| --- | --- | --- | --- | --- | --- |
|  |  | Disagree | Agree | Total |  |
| Age | Median | 33,00 | 29,00 | 29,00 | 0.236 |
|  | Percentile 25 | 23,00 | 21,00 | 22,00 |  |
|  | Percentile 75 | 44,00 | 38,00 | 38,00 |  |
| Post matric | Median | 9,00 | 6,00 | 6,00 | 0.200 |
|  | Percentile 25 | 5,00 | 4,00 | 4,00 |  |
|  | Percentile 75 | 14,00 | 11,00 | 11,00 |  |

*Table 13:* Associations between categorical demographic variables and vaccine confidence and belief in religious compatibility of vaccines

| Categorical demographic variables | | Vaccines are compatible with my religious beliefs | | | | | | p-value |
| --- | --- | --- | --- | --- | --- | --- | --- | --- |
|  |  | Disagree | | Agree | | Total | |  |
|  |  | Count | Row N % | Count | Row N % | Count | Row N % |  |
| Staff/Student | Staff | 12 | 5.2% | 217 | 94.8% | 229 | 100.0% | 0.352 |
|  | Student | 20 | 3.3% | 592 | 96.7% | 612 | 100.0% |  |
|  | Both | 4 | 5.1% | 75 | 94.9% | 79 | 100.0% |  |
|  | Total | 36 | 3.9% | 884 | 96.1% | 920 | 100.0% |  |
| Sex | Male | 12 | 5.2% | 220 | 94.8% | 232 | 100.0% | 0.234 |
|  | Female | 23 | 3.4% | 663 | 96.6% | 686 | 100.0% |  |
|  | Total | 35 | 3.8% | 883 | 96.2% | 918 | 100% |  |
| degree | BSc | 16 | 4.8% | 314 | 95.2% | 330 | 100.0% | 0.123 |
|  | Hons | 8 | 6.3% | 118 | 93.7% | 126 | 100.0% |  |
|  | MBBS | 5 | 2.5% | 197 | 97.5% | 202 | 100.0% |  |
|  | MSc | 7 | 3.9% | 174 | 96.1% | 181 | 100.0% |  |
|  | PhD | 0 | 0.0% | 81 | 100.0% | 81 | 100.0% |  |
|  | Total | 36 | 3.9% | 884 | 96.1% | 920 | 100.0% |  |
| Religion | Islam | 3 | 2.9% | 100 | 97.1% | 103 | 100.0% | 0.851 |
|  | Roman Catholic | 4 | 5.0% | 76 | 95.0% | 80 | 100.0% |  |
|  | Orthodox | 15 | 5.2% | 276 | 94.8% | 291 | 100.0% |  |
|  | Pentecostal | 7 | 4.0% | 169 | 96.0% | 176 | 100.0% |  |
|  | Traditional | 1 | 1.5% | 66 | 98.5% | 67 | 100.0% |  |
|  | Jewish | 0 | 0.0% | 7 | 100.0% | 7 | 100.0% |  |
|  | Buddhist | 0 | 0.0% | 3 | 100.0% | 3 | 100.0% |  |
|  | Hindu | 0 | 0.0% | 24 | 100.0% | 24 | 100.0% |  |
|  | Atheist | 4 | 5.3% | 71 | 94.7% | 75 | 100.0% |  |
|  | Agnostic | 2 | 3.0% | 64 | 97.0% | 66 | 100.0% |  |
|  | Other | 0 | 0.0% | 21 | 100.0% | 21 | 100.0% |  |
|  | 7th Day Adventist | 0 | 0.0% | 7 | 100.0% | 7 | 100.0% |  |
|  | Total | 36 | 3.9% | 884 | 96.1% | 920 | 100.0% |  |
| Age group | ≤24 | 13 | 3.5% | 363 | 96.5% | 376 | 100.0% | 0.756 |
|  | 25-34 | 7 | 3.2% | 213 | 96.8% | 220 | 100.0% |  |
|  | 35-44 | 7 | 4.0% | 166 | 96.0% | 173 | 100.0% |  |
|  | 45-54 | 4 | 5.3% | 71 | 94.7% | 75 | 100.0% |  |
|  | 55-64 | 4 | 6.2% | 61 | 93.8% | 65 | 100.0% |  |
|  | ≥65 | 1 | 9.1% | 10 | 90.9% | 11 | 100.0% |  |
|  | Total | 36 | 3.9% | 884 | 96.1% | 920 | 100.0% |  |

*Table 14:* The association between quantitative variables and belief in religious compatibility of vaccines

| Quantitative variables | | Vaccines are compatible with my religious beliefs | | | p-value |
| --- | --- | --- | --- | --- | --- |
|  |  | Disagree | Agree | Total |  |
| Age | Median | 33,00 | 29,00 | 29,00 | 0.267 |
|  | Percentile 25 | 22,00 | 22,00 | 22,00 |  |
|  | Percentile 75 | 43,50 | 38,00 | 38,00 |  |
| Post matric | Median | 5,50 | 6,00 | 6,00 | 0.614 |
|  | Percentile 25 | 3,00 | 4,00 | 4,00 |  |
|  | Percentile 75 | 12,50 | 11,00 | 11,00 |  |

*Table 15:* Associations between categorical demographic variables and intention to receive COVID 19 vaccine

| Categorical demographic variables | | I will take a Covid-19 vaccine when one becomes available | | | | | | p-value |
| --- | --- | --- | --- | --- | --- | --- | --- | --- |
|  |  | Disagree | | Agree | | Total | |  |
|  |  | Count | Row N % | Count | Row N % | Count | Row N % |  |
| Staff/Student | Staff | 19 | 8.3% | 210 | 91.7% | 229 | 100.0% | 0.383 |
|  | Student | 63 | 11.1% | 504 | 88.9% | 567 | 100.0% |  |
|  | Both | 10 | 13.0% | 67 | 87.0% | 77 | 100.0% |  |
|  | Total | 92 | 10.5% | 781 | 89.5% | 873 | 100.0% |  |
| Sex | Male | 20 | 8.5% | 216 | 91.5% | 236 | 100.0% | 0.317 |
|  | Female | 70 | 11.0% | 565 | 89.0% | 635 | 100.0% |  |
|  | Other | 2 | 100.0% | 0 | 0.0% | 2 | 100.0% |  |
|  | Total | 92 | 10.5% | 781 | 89.5% | 873 | 100.0% |  |
| degree | BSc | 35 | 11.7% | 263 | 88.3% | 298 | 100.0% | 0.161 |
|  | Hons | 18 | 16.1% | 94 | 83.9% | 112 | 100.0% |  |
|  | MBBS | 18 | 9.0% | 182 | 91.0% | 200 | 100.0% |  |
|  | MSc | 15 | 8.5% | 162 | 91.5% | 177 | 100.0% |  |
|  | PhD | 6 | 7.0% | 80 | 93.0% | 86 | 100.0% |  |
|  | Total | 92 | 10.5% | 781 | 89.5% | 873 | 100.0% |  |
| religion | Islam | 7 | 7.5% | 86 | 92.5% | 93 | 100.0% | . |
|  | Roman Catholic | 11 | 12.8% | 75 | 87.2% | 86 | 100.0% |  |
|  | Orthodox | 31 | 11.6% | 237 | 88.4% | 268 | 100.0% |  |
|  | Pentecostal | 22 | 13.7% | 139 | 86.3% | 161 | 100.0% |  |
|  | Traditional | 7 | 11.1% | 56 | 88.9% | 63 | 100.0% |  |
|  | Jewish | 1 | 14.3% | 6 | 85.7% | 7 | 100.0% |  |
|  | Buddhist | 1 | 25.0% | 3 | 75.0% | 4 | 100.0% |  |
|  | Hindu | 0 | 0.0% | 24 | 100.0% | 24 | 100.0% |  |
|  | Atheist | 4 | 5.6% | 68 | 94.4% | 72 | 100.0% |  |
|  | Agnostic | 2 | 2.9% | 66 | 97.1% | 68 | 100.0% |  |
|  | Other | 2 | 10.0% | 18 | 90.0% | 20 | 100.0% |  |
|  | 7th Day Adventist | 4 | 57.1% | 3 | 42.9% | 7 | 100.0% |  |
|  | Total | 92 | 10.5% | 781 | 89.5% | 873 | 100.0% |  |
| Age group | ≤24 | 38 | 10.9% | 311 | 89.1% | 349 | 100.0% | 0.992 |
|  | 25-34 | 20 | 9.8% | 185 | 90.2% | 205 | 100.0% |  |
|  | 35-44 | 18 | 10.8% | 148 | 89.2% | 166 | 100.0% |  |
|  | 45-54 | 7 | 9.3% | 68 | 90.7% | 75 | 100.0% |  |
|  | 55-64 | 8 | 11.9% | 59 | 88.1% | 67 | 100.0% |  |
|  | ≥65 | 1 | 9.1% | 10 | 90.9% | 11 | 100.0% |  |
|  | Total | 92 | 10.5% | 781 | 89.5% | 873 | 100.0% |  |

*Table 16:* Associations between quantitative variables and intention to receive COVID 19 vaccine

| Quantitative variables | | I will take a Covid-19 vaccine when one becomes available | | | p-value |
| --- | --- | --- | --- | --- | --- |
|  |  | Disagree | Agree | Total |  |
| Age | Median | 30,00 | 30,00 | 30,00 | 0.990 |
|  | Percentile 25 | 21,00 | 22,00 | 22,00 |  |
|  | Percentile 75 | 38,00 | 39,00 | 38,00 |  |
| Post matric | Median | 5,00 | 6,00 | 6,00 | 0.301 |
|  | Percentile 25 | 3,00 | 4,00 | 4,00 |  |
|  | Percentile 75 | 12,00 | 11,00 | 11,00 |  |

*Table 17a:* Association between levels of education, vaccine confidence statements and the intention to receive a COVID 19 vaccine

| Statements | Undergraduates  (BSc. & MBBS) | | Postgraduates  (Honours & above) | | Pearson’s chi square 2-sided p-value |
| --- | --- | --- | --- | --- | --- |
|  | Agree | Disagree | Agree | Disagree | 0.841 |
| 1. Vaccines are important for children to have | 92.2% | 2.8% | 97.6% | 2.4% |  |
| 2. Vaccines are important for me to have | 95.1% | 4.9% | 95.9% | 4.1% | 0.664 |
| 3. Overall, I think vaccines are safe | 95.9% | 4.1% | 94.7% | 5.3% | 0.431 |
| 4. Overall, I think vaccines are effective | 98.0% | 2.0% | 96.6% | 3.4% | 0.218 |
| 5. Vaccines are compatible with my religious beliefs | 96.1% | 3.9% | 96.1% | 96.1% | 1.000 |
| 6. I will take a Covid-19 vaccine when one becomes available | 89.4% | 10.6% | 89.6% | 10.4% | 1.000 |

| Statements | Up to Honors  (BSc., MBBS, & Honors) | | Masters & above  (Masters & Ph.D.) | | p-value |
| --- | --- | --- | --- | --- | --- |
|  | Agree | Disagree | Agree | Disagree | 0.381 |
| 1. Vaccines are important for children to have | 97.0% | 3.0% | 98.3% | 1.7% |  |
| 2. Vaccines are important for me to have | 94.5% | 5.5% | 97.6% | 2.4% | 0.043 |
| 3. Overall, I think vaccines are safe | 95.2% | 4.8% | 96.0% | 4.6% | 0.612 |
| 4. Overall, I think vaccines are effective | 97.2% | 2.8% | 97.9% | 2.1% | 0.660 |
| 5. Vaccines are compatible with my religious beliefs | 95.6% | 4.4% | 97.3% | 2.7% | 0.262 |
| 6. I will take a Covid-19 vaccine when one becomes available | 88.4% | 11.6% | 92.0% | 8.0% | 0.119 |

Table17b: Association between levels of education, vaccine confidence statements and the intention to receive a COVID 19
